# Can medication mitigate the need for a strict lock down?: A mathematical study of control strategies for COVID-19 infection

**DOI:** 10.1101/2020.05.29.20116749

**Authors:** Mohsin Ali, Mudassar Imran, Adnan Khan

**Author notes:** Author email: mohsin (Mohsin Ali).

## Abstract

We formulate a deterministic epidemic model to study the effects of medication on the transmission dynamics of Corona Virus Disease (COVID-19). We are especially interested in how the availability of medication could change the necessary quarantine measures for effective control of the disease. We model the transmission by extending the SEIR model to include asymptomatic, quarantined, isolated and medicated population compartments. We calculate the basic reproduction number *R*_0_ and show that for *R*_0_ < 1 the disease dies out and for *R*_0_ > 1 the disease is endemic. Using sensitivity analysis we establish that *R*_0_ is most sensitive to the rates of quarantine and medication. We also study how the effectiveness and the rate of medication along with the quarantine rate affect *R*_0_. We devise optimal quarantine, medication and isolation strategies, noting that availability of medication reduces the duration and severity of the lock-down needed for effective disease control. Our study also reinforces the idea that with the availability of medication, while the severity of the lock downs can be eased over time some social distancing protocols need to be observed, at least till a vaccine is found. We also analyze the COVID-109 outbreak data for four different countries, in two of these, India and Pakistan the curve is still rising, and in he other two, Italy and Spain, the epidemic curve is now falling due to effective quarantine measures. We provide estimates of *R*_0_ and the proportion of asymptomatic individuals in the population for these countries.

## 1 Introduction

Originating in Wuhan, China, with the first reported cases in early December 2019[2], Coronavirus disease (COVID-19) is a pandemic that has spread around the globe in the early months of 2020. The disease is prevalent in at least 212 countries and territories, with more than 5.3 million cases reported and around 325,000 fatalities at the time of writing on May 22, 2020 [3].

The causative agent of COVID-19 is a coronavirus SARS-CoV-2 [2]. This belongs to a family of viruses that are found in humans and different species of animals including cattle, camels and bats. These have in the past caused serious disease outbreaks in human populations as happened in MERS (caused by MERS-CoV) and SARS (caused by SARS-CoV) [1, 4, 5], it has been reported that both these viruses like SARS-CoV-2 have their origins in bats.

The virus is mainly spread from person to person, through respiratory droplets, the spread is more likely when people are within 6 feet of each other[1]. Symptoms of the disease may appear 2-14 days after exposure and may include fever, cough shortness of breath, chills, muscle pain and loss of taste or smell[2, 6]. Many of these are common with influenza, however high persistent fever, dry cough and difficulty in breathing [6] seem to characterize COVID-19. The symptoms may range from very mild (in 80 % of the cases) to severe (in 15 % of the cases) to critical (in 5 % of the cases)[2]. Those at higher risk for severe illness include the elderly (people ages 65 and above) and people with underlying medical conditions which may include chronic lung diseases, diabetes, chronic kidney diseases, serious heart conditions and immunocompromised individuals[14].

At this time there is no available vaccine for COVID-19, and the available antiviral treatments are all in the trial phase[1]. This prompted health authorities to stress upon social distancing as a means to control the spread of the disease [2]. Lock downs were enforced in various countries, however these measures have resulted in substantial economic and social cost [7]. As a consequence many countries have started to ease the lock downs, despite concerns expressed by public health practitioners. There may be some positive news as in some recent studies antiviral treatments have shown some promising results, reducing the infectious period and alleviating disease symptoms [9, 8, 12, 13].

As the disease burden of COVID-19 continues to rise steeply, there is a rush to verify the benefits of different treatment regimens for which there is any anecdotal evidence available. Initial clinical trials so far have produced mixed results with some drug regimens showing encouraging outcomes, such as shortening of the duration of the disease, while others showing no significant change in the outcomes due to medication. The anti malarial hydroxychloroquine has been suggested as possible medication for COVID-19. While the National Institute of Health (NIH) in the United States has began clinical trials for hydroxychloroquine taken together with azithromycin [15], a recent study by Mehra et.al [10] has been unable to confirm any benefit of hydroxychloroquine when used alone or with a macrolide (such as Azithromycin), on in-hospital outcomes for COVID-19, in fact, each of these drug regimens was associated with decreased in-hospital survival. Another drug that has been suggested to have beneficial effects in COVID-19 treatment is Remdesivir. Some initial studies on the effectiveness of Remdesivir have been published; Wang et.al [11] reported no significant clinical improvement in COVID-19 symptoms using the drug, they do recommend further clinical studies be done, however Grein et.al [13] have reported improvement in 68% of the patients in a controlled trial using the drug, similarly Beigel et.al [9] have also reported shortening in the time to recovery with Remdesivir use. Huang et.al [6] used a triple anti viral therapy in their study and have reported alleviation of symptoms and shortening the duration of the disease. At the time of writing several studies are being carried out for a variety of plausible treatments which have shown some positive results. Based on this one can be cautiously optimistic about the possibility that medication that alleviates symptoms and shortens the duration of the disease may be available soon.

Since the outbreak many mathematical models have been proposed to estimate the growth rates and understand the transmission dynamics of COVID-19. These include phenomenological models[16, 17], stochastic models[18], both of which are very useful in the early stages of the outbreak, and mechanistic models[19, 20, 22, 23, 30] that incorporate our understanding of the transmission pathways. Imran et.al [24] and Perkins et.al [31] have used optimal control techniques to propose efficient control strategies. The aim of such modeling is twofold, one to provide estimates of the severity of the outbreak by calculating quantities like the growth trends of the epidemic, estimates of the final outbreak size and duration of the outbreak and second to provide insights into efficacy of various control measures.

In the literature a variety of mathematical models have been proposed to study the role of medication in controlling different epidemics. Lee et.al [25] consider an extension of the SEIR model with isolation and treatment to study various treatment and isolation strategies for controlling an influenza pandemic. Granich et.al [27] use an SIR model with multiple infection and treatment stages to study the effects of antiretroviral therapy and universal voluntary reporting for HIV. Jia et.al [26] study the effectiveness of various control strategies including treatment using an SEIR model with compartments for acute and chronic stages of infection and treatment. Sharomi et.al [28] use an extension for the SEIR model to study HIV/TB co-infection and do a cost/benefit analysis of various treatment strategies. Tufail et.al [29] study a model for the transmission ad control of the H1N1 Influenza (Swine Flu) using an extended SEIR model with treatment, vaccination and hospitalization compartments.

This study is motivated by some promising reports about the success of various drug therapies in medical trials as discussed above. We are interested in studying the effects of medication on the transmission dynamics of COVID-19 and in particular the effects on quarantine and other control strategies. We extend an earlier model by Imran et.al [24] to model the transmission dynamics of COVID-19. The model incorporates asymptomatic, isolation and quarantine compartments, these are considered particularly relevant in the transmission of COVID-19, we also include a compartment representing the medicated population group. We establish a threshold quantity, the basic reproductive number *R*_0_, the disease dies out if *R*_0_ < 1 and is endemic in the population when *R*_0_ > 1. We calculate the sensitivity of *R*_0_ on various model parameters, identifying the contact rate, the rates of entering and leaving the quarantine and the rate and efficacy of the medication,as parameters to which *R*_0_ is most sensitive. We also study the variation in *R*_0_ as the quarantine rate and the rate and the efficacy of the medication is varied, leading to some interesting observations. Using techniques from optimal control theory, we propose efficient quarantine, isolation and medication strategies. We also compare the quarantine and isolation strategies with and without the availability of medication, which leads to some insights regarding easing of lock downs once medication is available. Finally, we estimate *R*_0_ for different countries, which are at different phases of the epidemic. In India and Pakistan the epidemic is still in the growth phase, the dynamics of the disease also seem to be different here than in Europe and North America, with a much slower growth and mortality rate. We also fit the our model to the outbreak data to estimate the proportion of asymptomatics in the population and the average quarantine rate in these countries.

## 2 Model Formulation

The COVID-19 transmission model we consider is based upon the SEIR model and takes into account the effects of quarantine, isolation, medication and asymptomatic individuals. The total population *N*(*t*) is divided into ten mutually exclusive sub populations, susceptibles *S*, these are individuals who can fall ill by coming in contact with an infected individual, quarantined susceptibles *Q_S_*, these individuals are removed from the susceptible group at rate *ϵ*, either through self quarantined or lock-down measures, they however go back to the susceptible group at rate *ξ*. The susceptibles move to the exposed class by coming in contact with any infectious individual, at rate *ρλ*, some exposed individuals will not show symptoms and are accounted for in the model by the movement to the asymptomatic class at rate (1 − *ρ*)*λ*. Due to lock down measures, the exposed and asymptomatic sub-populations are also quarantined at rate *κ_E_* and *κ_A_*, respectively. The exposed and exposed quarantined individuals become infected at rates *σ* and *ν*, while some of the quarantined exposed are also isolated at the rate of *ω_E_*. Infected individuals can be isolated or given medication at a rate of *τ* and *α_I_* whereas a fraction of isolated are given medication at rate *α_Q_*. The recovery time for infected, asymptomatic, asymptomatic quarantined, isolation and medication is given by 1/γ,1/*θ_A_*,1/*ω_A_*,1/*θ* and 1/*θ_M_* respectively. The schematic of the transmission pathways is given in Fig.1. The total population *N*(*t*) is given by the sum of the sub-populations.

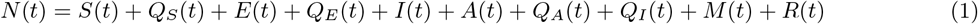

**Figure 1:**
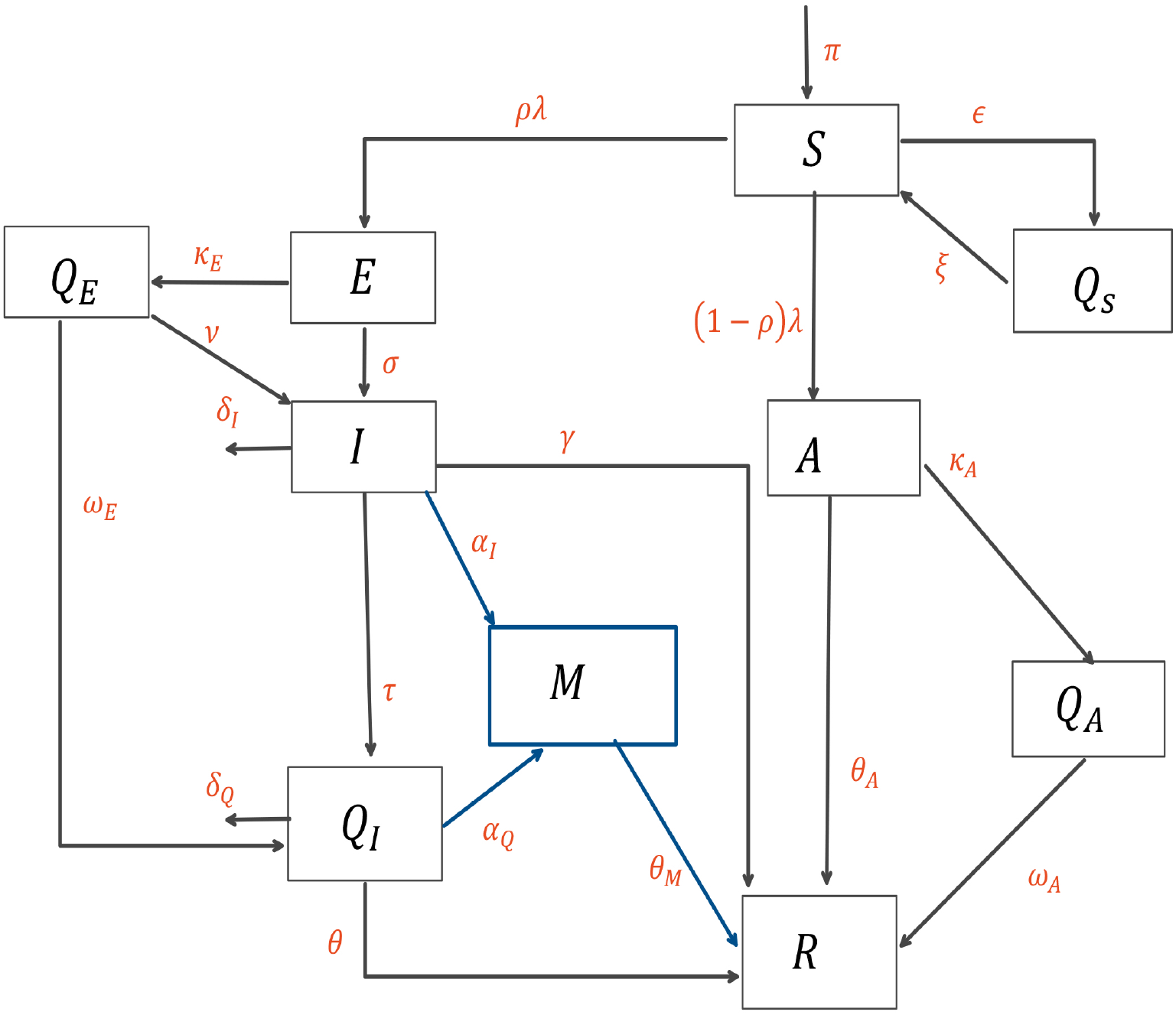
Schematic diagram of the model (2)

The governing equations are given below (2).

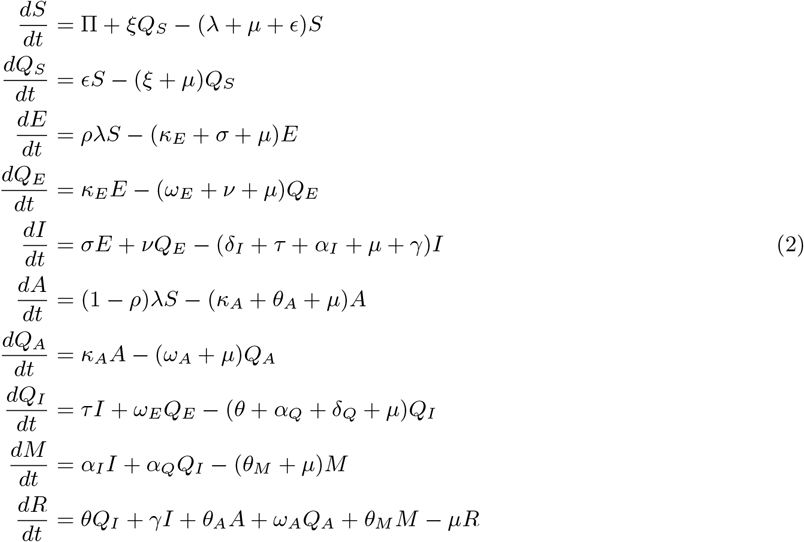

where *λ* is the force of infection

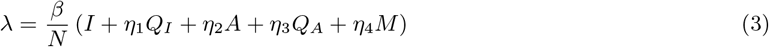

Here *β* is the effective contact rate, where as *η*_1_,*η*_2_ and *η*_3_ are the associated relative infectiousness parameters for the *Q_I_*,*A*, *Q_A_* and *M* sub-populations. Table.3 and Table.4 represent the description of variables and parameters of the model, and are given in the appendix.

## 3 Basic Properties

The described state variables in model (2) display non-negative solutions for all time *t* ≥ 0 for non-negative initial conditions.

### Lemma 3.1

*For any given non-negative initial conditions, there exist a unique solution S*, *Q_S_*, *E*, *Q_E_*, *I*, *A, Q_A_, Q_I_*, *M*, *R respectively, for all t ≥* 0. *Moreover, it satisfy the following inequality of boundedness*.

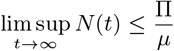

Proof is presented in the appendix A.

### Lemma 3.2

The closed set:

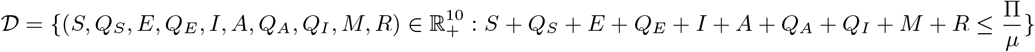

*is positively invariant*.

Proof is attached in the appendix A.

## 4 Steady State Analysis

### 4.1 Disease Free Equilibrium

The transmission model (2) attains the Disease Free Equilibrium (DFE) whenever there is no infection induced by the disease i.e. the force of infection is zero, *λ* = 0. Mathematically we can find this equilibrium state by equating the right hand side of (2) to zero with zero force of infection. Let 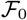 represent the DFE of the model.

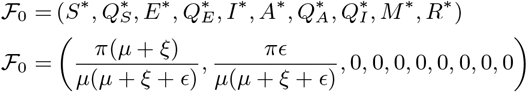

The local stability of this steady state is governed by the threshold quantity 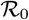 which is obtained using the next generation operator method [32].

### 4.2 The Basic Reproduction Number ℛ_0_

The thresh hold quantity 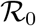 represents the average number of secondary infections generated by the single infection in the completely susceptible population. It is determined by the spectral radius of the *FV^−^*^1^ matrix. [32]. The corresponding *F* and *V* matrices of model (2) are given as

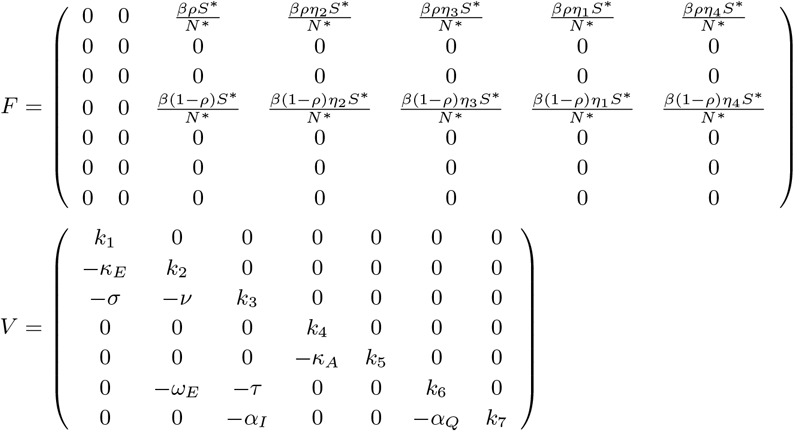

where *k*_1_ = *κ_E_* + *σ* + *μ*, *k*_2_ = *ω_E_* + *ν* + *μ*, *k*_3_ = *δ_I_* + *τ* + γ + *α_I_* + *μ*, *k*_4_ = *κ_A_ + θ_A_ + μ, k_5_ = ω_A_ + μ, k*_6_ *= θ + δQ + α_Q_ + μ, k*_7_ *= θ_M_ + μ*

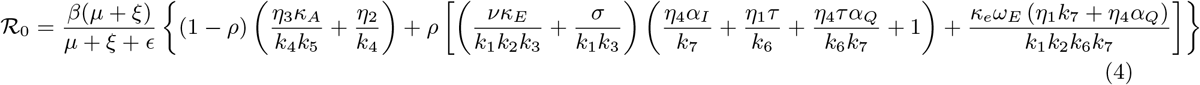

#### Stability of DFE

##### Lemma 4.1

*The steady state* (*DFE*) 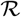 *of the model* (2) *is locally-asymptotically stable if* 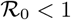, *and unstable if* 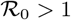.

The basic interpretation of this lemma is that a small influx of the infected individuals will not cause large outbreaks and disease will become extinct in long run.

To verify that the disease extinction is not dependent on the initially available sub-populations in model (2), it is necessary to show that the steady state (DFE) is globally asymptotically stable (GAS).

##### Lemma 4.2

*The Steady state* (*DFE*) 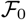 *of model is globally asymptotically stable if* 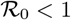 *and unstable* 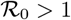. Proof is given in appendix A

### 4.3 Endemic Equilibrium

The steady state of the system (2) in presence of the infection i.e. *λ* ≠ 0 is known as the endemic equilibrium. Let 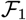 represents the arbitrary endemic equilibrium of the model (2).

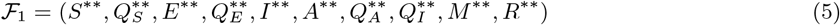

Further, The force of infection λ can be written in terms of the equilibrium as

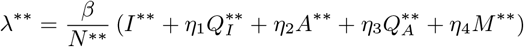

with 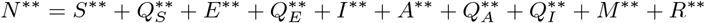

Solving for the system (2) at this specific fixed point, the endemic equilibrium becomes

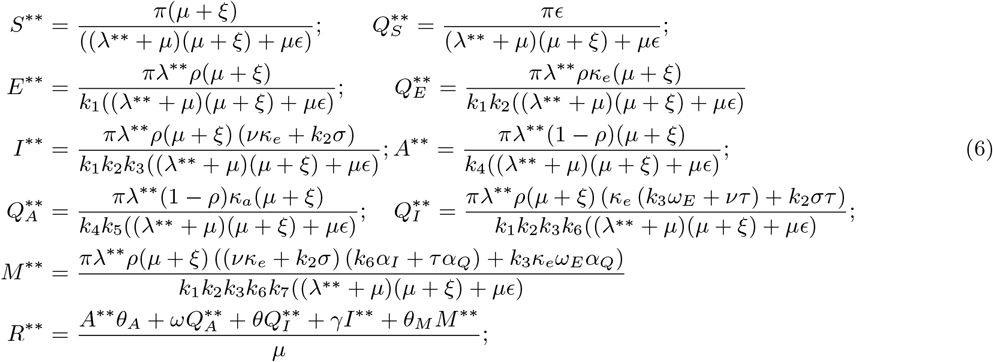

#### Lemma 4.3

*The model* (2) *achieves the unique positive endemic equilibrium whenever* 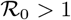.

#### Final size of disease COVID-19

In this section, we formulate the final size of the COVID-19 is estimated using the model (2) transmission. Applying the formulation by Arino et.al[34], define *u* ∊ ℝ*^n^*, 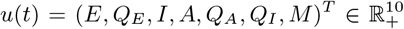 represents the set of infected compartments. Let *v ∊* ℝ*^m^*, *v*(*t*) = (*S*) ∊ ℝ_+_ and *w ∊* ℝ*^k^*, 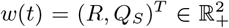 represents the susceptible and set of recovered and susceptible quarantine respectively. The model (2) can be restructured as

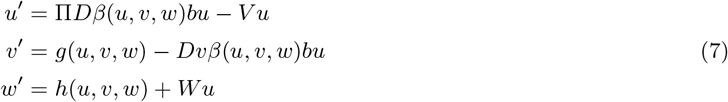

here *D* is *m* × *m* diagonal matrix with entries that are relative susceptibilities of *v* compartments. Π is *n* × *m* matrix with spacial property that (*i, j*)th entry represents the fraction of *j*th susceptible compartment goes to *i*th infected compartment. *b* is a *n* dimensional row vector with relative horizontal transmission coefficients. *g*(*u, v, w*) and *h*(*u, v, w*) are functions with uninfected individuals by means of birth, death, quarantine and recovery through natural immunity.

##### Theorem 4.4

*The final size of the epidemic in model* (2) *is given by*

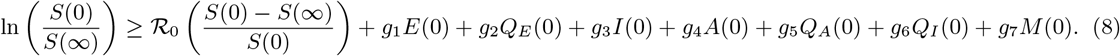

*where*

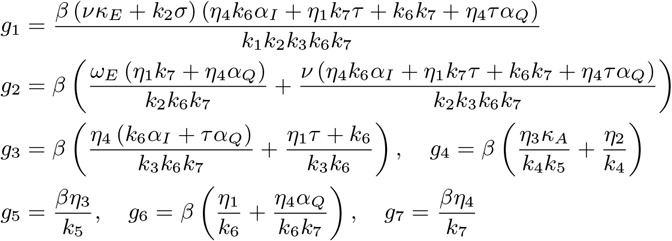

#### 4.3.1 Uniform Persistence

Let 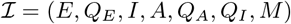. Then 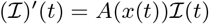, where

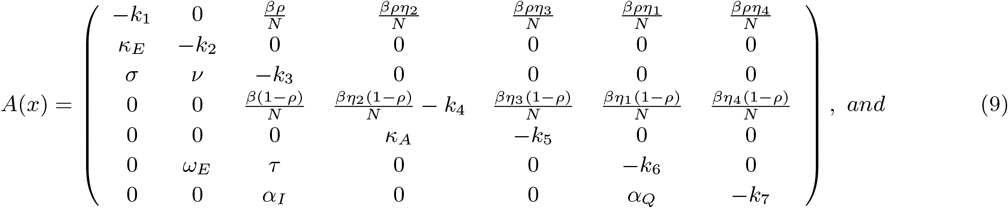

Denote by *s*(*A*) the spectral bound of matrix *A*. Let 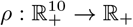

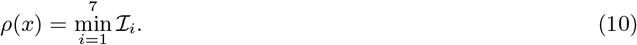

##### Theorem 4.5

*If R_0_ > 1 then the disease is strongly uniformly ρ-persistent: ∃ε > 0 such that*

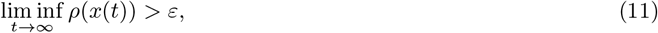

*whenever ρ*(*x*(*0*)) *> 0. Where x*(*t*) *=* (*S*(*t*)*, Q_S_*(*t*)*, E*(*t*)*, Q_E_*(*t*)*, I*(*t*)*;A*(*t*)*, Q_A_*(*t*)*, Q_I_* (*t*)*, M*(*t*)*, R*(*t*)) *be a solution of model* (*2*).

The above persistence result shows that the disease will persist and there will be an endemic steady state if the value of the basic reproductive number is greater than 1.

## 5 Numerical Simulations

Matlab(ODE 45) is used to perform the simulations with the parameter values given in the Tab.4. The time series solutions are are shown in Fig. 2, it is apparent that model (2) achieves the Endemic equilibrium respectively whenever the threshold quantity 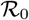 is more than the unity and the DFE whenever 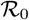 is less than the unity authenticating the qualitative results.

**Figure 2:**
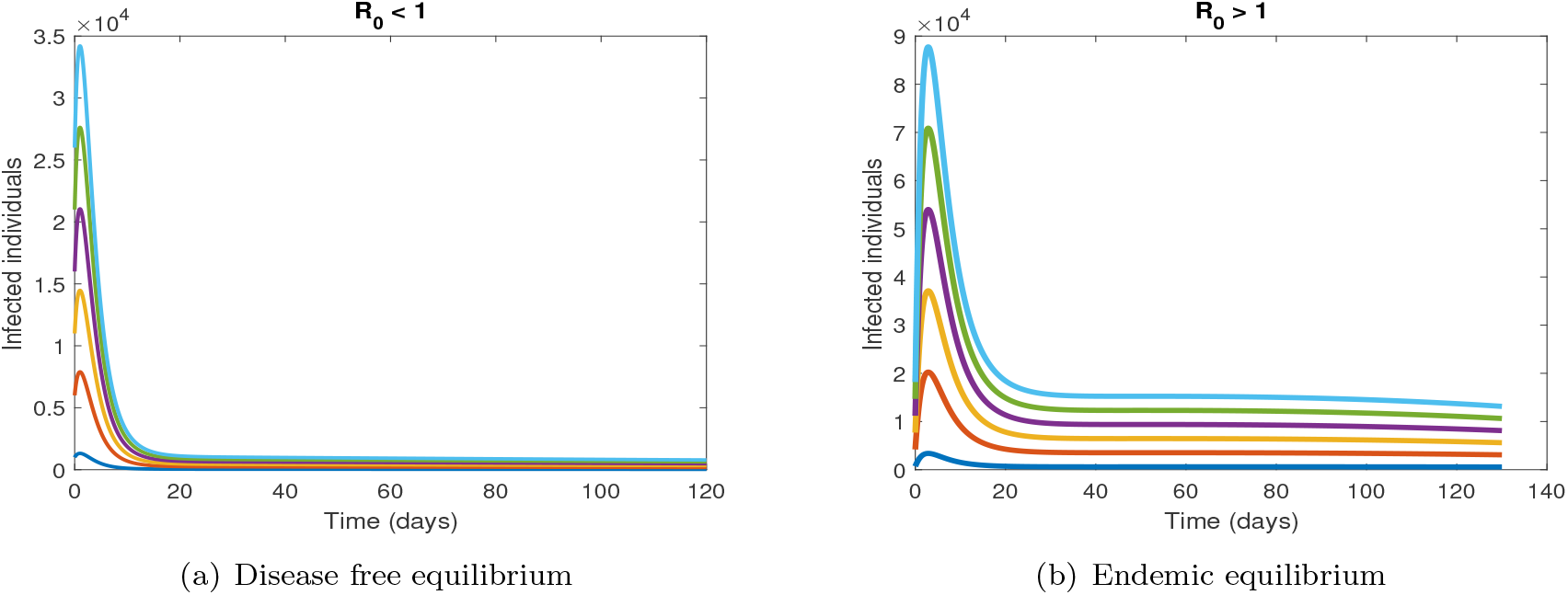
Time Series Analysis

We now investigate the effects of medication on the quarantined population, taking into account both the rate and efficacy of medication in Fig.3(a) and (c). We also look into the effect of the medication rate and efficacy on the number of days spent in quarantine in Fig.3(e) and (g).

**Figure 3:**
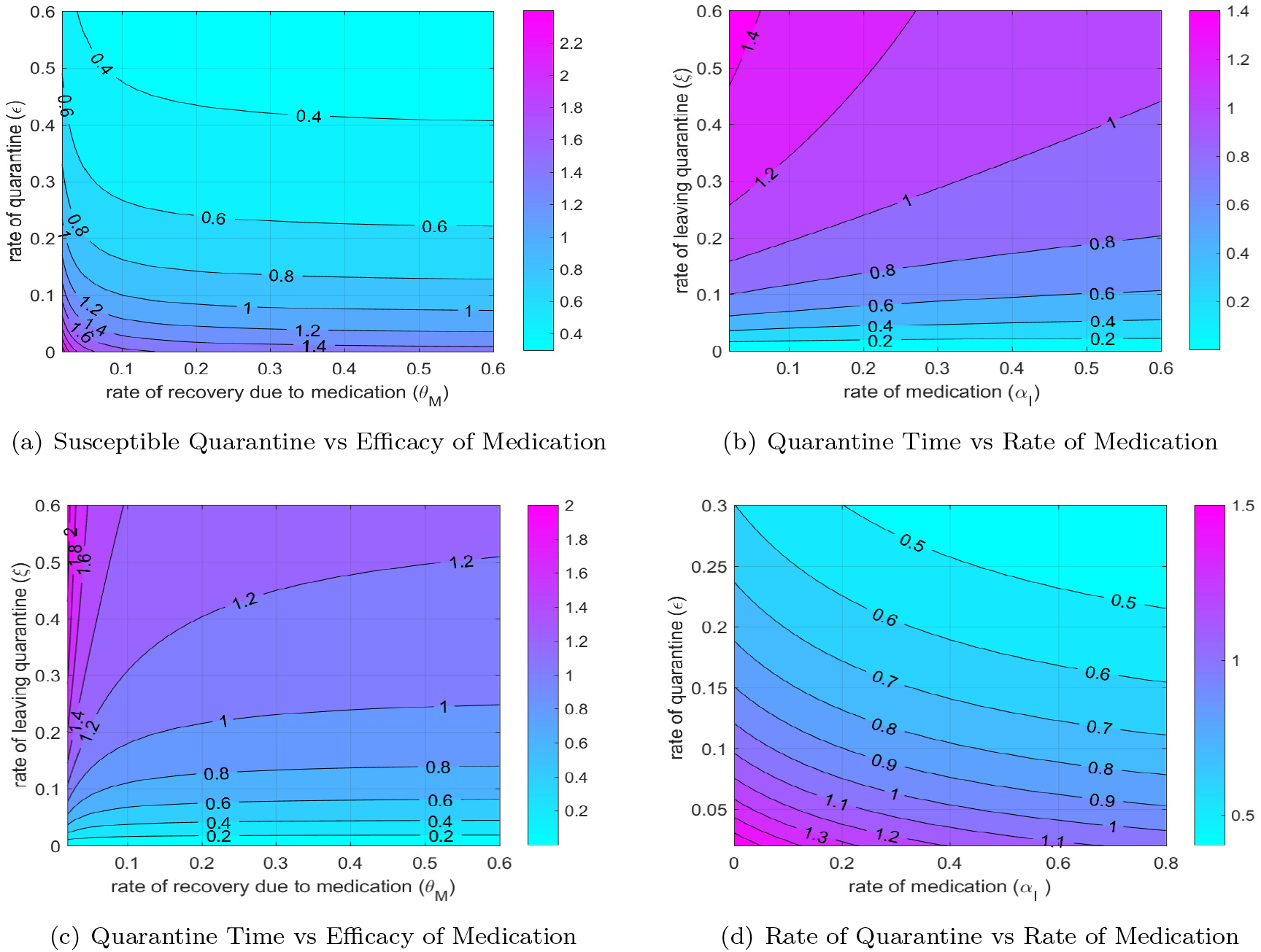
Contour’s of 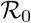

**Figure 4:**
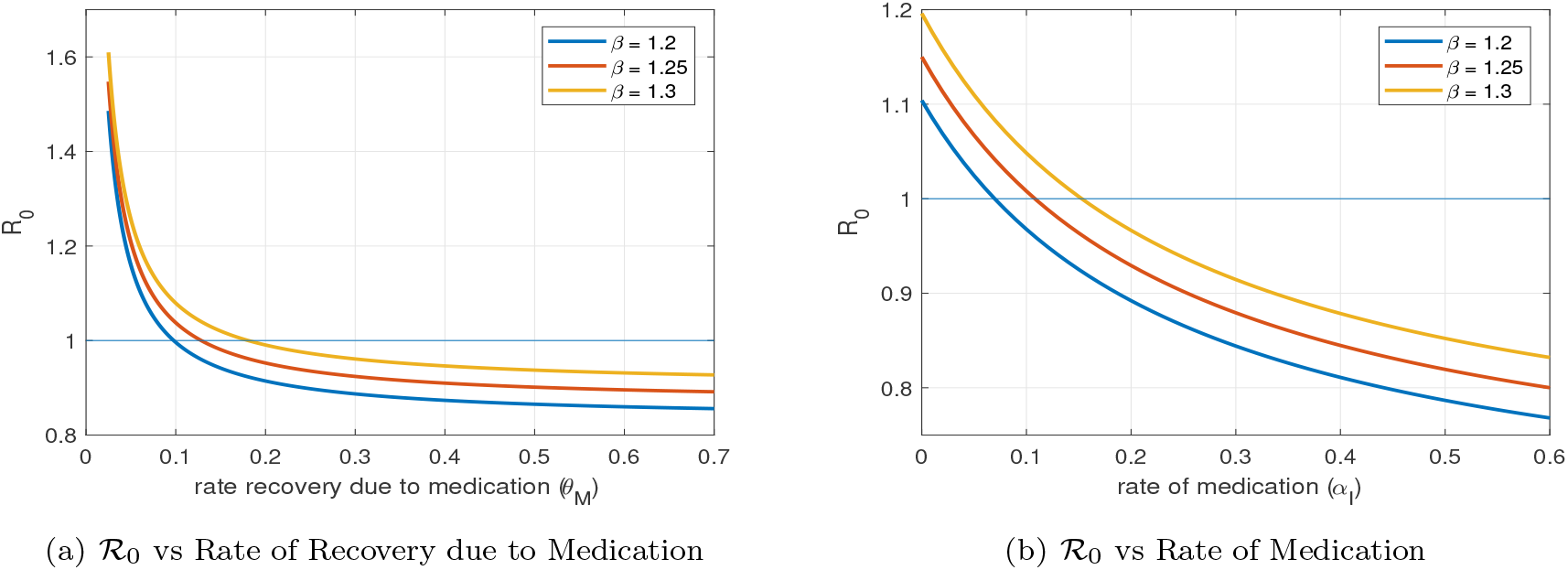
Dependence of 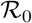 on Medication

We observe that for a realistic recovery rate due to medication (based on the studies thus far), we would have to maintain a quarantine rate of (20 %) or higher in order to have over thresh hold quantity 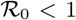, however for high quarantine rates a less effective medication will suffice. This illustrates that fact that in order to ease the lock down we need effective medication. Moreover,over a range of different realistic contact rates *β*,we need certain level of effectiveness of the medication as is shown in Fig.3(b).

These observations point towards the fact that even with the availability of medication, while the social distancing measures may be relaxed but they probably cannot be completely done away with until a vaccine is found.

## 6 Sensitivity Analysis

The source of sensitivity is due the variation found in the parameters. This section deals with the sensitive parameters to 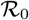 i.e. how much 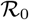 changes upon small perturbations of the parameters. This will gives us insights about the parameters that we can use in devising our control strategies. PRCC (Partial Rank Correlation Coefficient) method is effective tool which compute the impact of parameters on 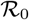. The positive(negative) PRCC indicates the positive(negative) correlation parameters with the output variable.The parameters involved in 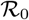 are studied which are *μ, ν, β, σ, τ, κ_E_, ω_E_*, *γ*, *θ_A_*, *κ_A_*, *ρ*, *∊*, *ξ*, *θ*, *θ_M_, α_I_, α_Q_*. Fig. 5 indicates that the most sensitive parameter of model (2) are *β*, *∊*, *τ* and *α_I_* thus pointing towards the control strategies such as quarantine, isolation and medication.

**Figure 5:**
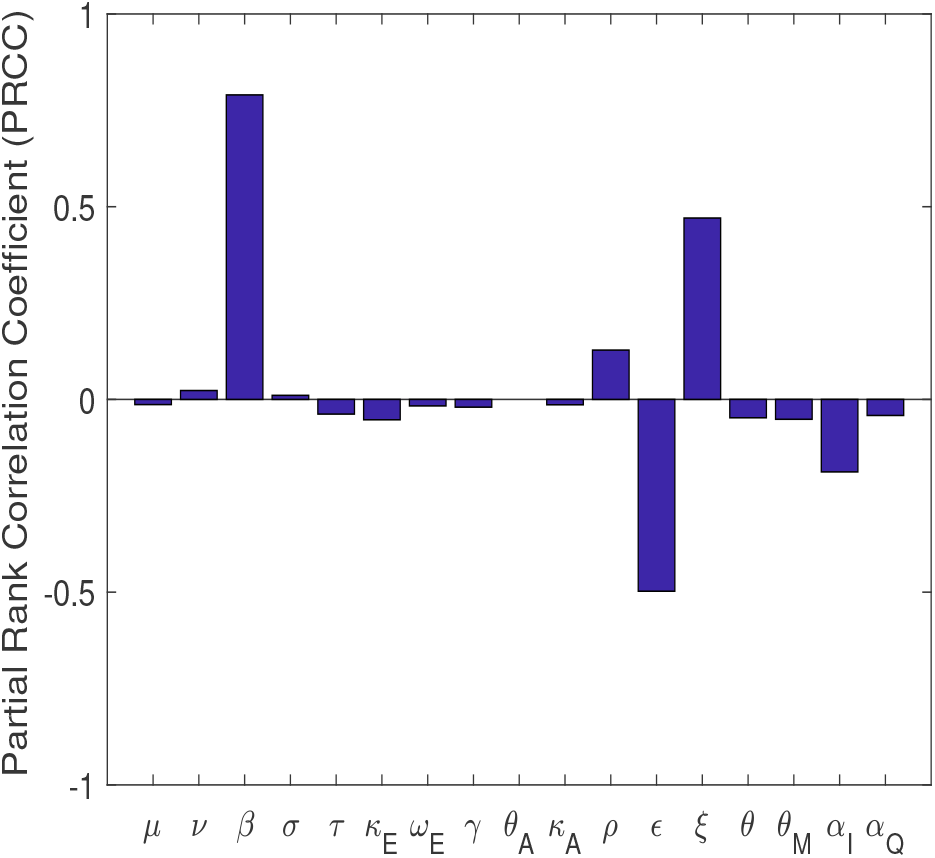
Sensitivity of the Parameters

## 7 Estimation of Basic Reproduction Number ℛ_0_

The thresh hold quantity 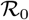, basic reproduction number is the expected number of the newly infected individuals caused by the single infected individual. It is convenient to estimate the 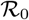 from the data in order to have better insights about COVID-19. The Estimates of R0 depend upon the estimation of critical parameters such as contact rate *β*, incubation rate *σ*, the recovery *γ*, and other involving parameters. The estimation is performed by fitting COVID-19 data of Italy, Spain, India, and Pakistan via nonlinear least square method. The underlying fitting model is based upon an extension of SEIR by incorporating the effect of Quarantine, Isolation, and Asymptomatic (the reduced model of Imran et al [24]).

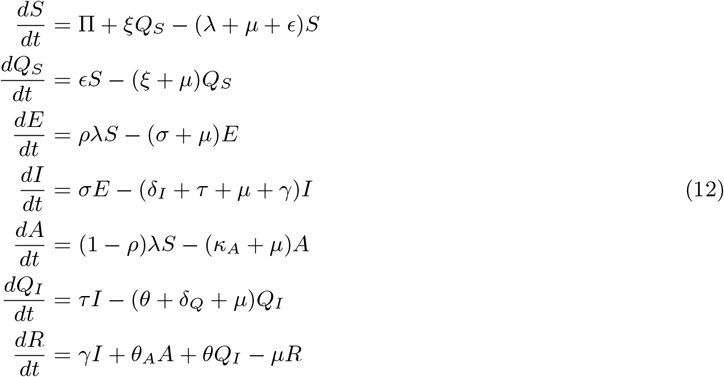

where *λ* is the force of infection

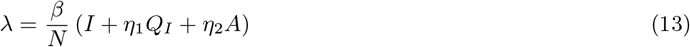

Here Π is the recruitment rate, which depends upon the demographic area from the data is taken. 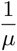 is the average life span of human life. The effective contact rate is the most crucial parameter, which describes the average associated chance of getting infected by the interaction of the susceptible individuals with the infected ones. The direct measure of beta is tricky unless an epidemic is completed. However, an indirect approach indicated by Chavez et al. [42] can be adopted to estimate the *β* for 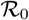.

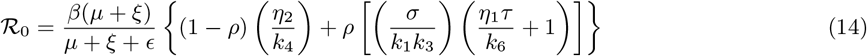

**Figure 6:**
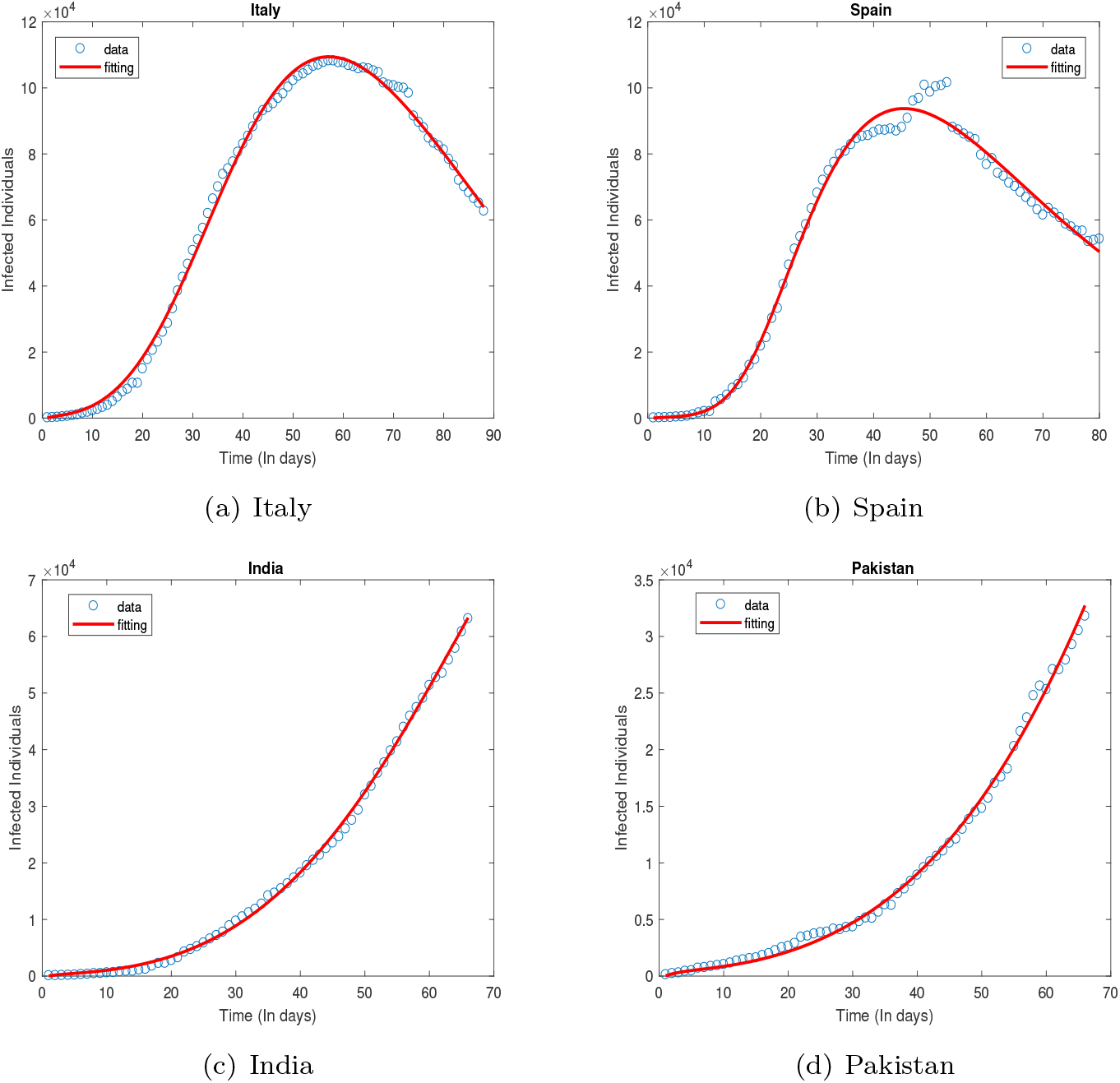
Parameter Estimation

The ordinary least squares (OLS) scheme is used to estimate the Tab 7 paramters. Since the observed data values is assumed to have constant variance error distribution. So, we can write the estimation scheme as

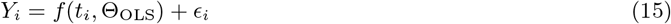

where *Y_i_* and Θ*_OLS_* are data observes and set of parameters involving fitting process respectively.

The optimized set of parameters can be obtained by minimizing the least squared sum over the set of estimated parameters.

The Matlab tool is used to perform this optimization under the trust-region-reflective algorithm to find the best fit of the selected parameters. The bootstrap method suggested by the Efron and Tibshirani [43] to use the associated 95 % confidence intervals.

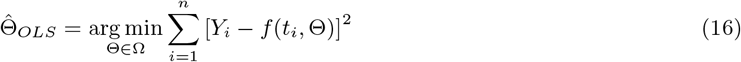

**Table 1:** Parameter Estimates for Different Countries

| Parameters | Italy | Spain | India | Pakistan |
| --- | --- | --- | --- | --- |
| $\beta$ | 1.4 (1.3-1.4) | 2.2 (2.1-2.4) | 0.87 (0.85-0.91) | 0.68 (0.66-0.71) |
| $\sigma$ | 0.33 (0.31-0.35) | 0.11 (0.11-0.12) | 0.19 (0.19-0.2) | 0.15 (0.15-0.16) |
| $\gamma$ | 0.099 (0.094-0.1) | 0.15 (0.14-0.17) | 0.12 (0.11-0.14) | 0.11 (0.10-0.12) |
| $\epsilon$ | 0.11 (0.10,0.12) | 0.054 (0.05-0.066) | 0.079 (0.074-0.082) | 0.065 (0.062-0.068) |
| $\rho$ | 0.8 (0.79-0.81) | 0.74 (0.73-0.75) | 0.72 (0.71 – 0.73) | 0.66 (0.65-0.68) |
| $\mathcal{R}_0$ | 1.1408 | 2.6327 | 1.3144 | 1.2303 |

**Table 2:** Estimates of 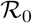 for Different Epidemic Phases

| $\mathcal{R}_0$ | Italy | Spain | India | Pakistan |
| --- | --- | --- | --- | --- |
| Initial Phase | 2.15 | 2.59 | 1.3144 | 1.2303 |
| Up to Middle Phase | 2.99 | 3.7 | - | - |
| Up to Now | 1.1408 | 2.6327 | 1.3144 | 1.2303 |

We note that the values of 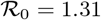 and 1.23 respectively for India and Pakistan are relatively small considering that the outbreak there is still in the growth phase. There has been a lot of speculation regarding why the outbreak in these countries has been less severe, but nothing conclusive can be said. The low value of the contact rate could mean either an effective quarantine or the probability of infection being lower. We also note that the proportion of asymptomatic individuals is 72% and 66% respectively in India and Pakistan, this is consistent with other estimates for a wider population as well.

For Italy where the epidemic curve is falling due to the strict lock down, we note that the value of *R*_0_ comes out to be 1.14, similarly for Spain *R*_0_ comes out to be 2.6, the values seems to be on the higher side and can be attributed to a sharp increase in the number of cases around week 7.The asymptomatic population is estimated to be 80% an 74% for Italy and Spain, again within the range of estimates in the literature.

## 8 Control Strategies for the Outbreak

The theory of optimal control developed by Lev Pontryagin and his collaborators is used for models where the underlying dynamics are governed by systems of differential equations. The ‘Pontryagin’s maximum principle’ algorithm allows us to minimize a ‘cost functional’ subject to differential equation constraints. It has found wide application in biological models including epidemic models [25, 26, 29]. The goal here is to reduce the infected population by means of specific controls, which may appear as time dependent parameters in the model, while minimizing the required resources. The algorithm is implemented by appending an adjoint system of differential equations having a terminal conditions along with the original state system. Further details regarding Optimal Control and adjoint system can be found in [36], [37]. In our study numerical results are produced using the forward (state system) backward (adjoint system) sweep method with a fourth-order backward Runge-Kutta method.

In this paper, we we devise optimal control strategies for quarantine of susceptibles and the isolation and medication of infected individuals that will reduce the infection while keeping the cost at a minimum. The cost includes the costs associated with the disease burden, as well as the cost of implementing the control strategy.

### 8.1 Optimal Medication with Isolation and Quarantine

In the recent COVID-19 pandemic, the most effective and widely used method to reduce the infection has been the quarantine of the susceptible population, along with isolation of the confirmed infected individuals. We now have the possibility of having medication available to help control the outbreak as mentioned in section 1 Introduction. In this section, we determine time dependent strategies for the medication rate *α_I_*, isolation rate *τ* and the rate of quarantine *∊* to effectively control the infected population and at the same time keeping the associated cost low. Let *U* be the control set defined for the three parameters *a_I_*, t, e

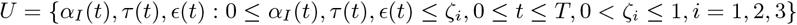

here *α_I_*(*t*)*,τ*(*t*), *ϵ*(*t*) are Lebesgue measurable and *ζ_i_*, ∀*i* = 1, 2, 3 are positive numbers which are maximum values for the respective parameters. Our main objective to minimize the functional involving the infected sub populations along with the described control parameters.

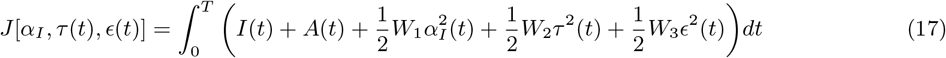

where *W_i_* for *i* = 1, 2, 3 are cost balancing coefficients assigned to the medication,isolation and susceptible quarantine parameters, these are needed as the parameters themselves have values between 0 and 1, while the infected population is in the thousands, further they can be used to assign relative importance to the different control parameters. Our focus is to find the optimal values for the control parameters 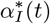, *τ*\*(*t*), *∊*\*(*t*) such that

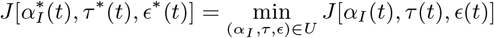

We are interested in studying the effects that medication may have on quarantine and isolation strategies. With the rising socio economic cost of a strict quarantine and the burden on health facilities to isolate infected individuals effective medication may alleviate some of the factors. We would also like to look into the costs associated with using medication along with quarantine and isolation to control the epidemic.

Our main result for this section is given below. For the details of formulation of the Hamiltonian and the adjoint system along with finding the optimality conditions the reader is referred to the Appendix.

#### Theorem 8.1

*There exist unique optimal controls* 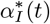, *τ*\*(*t*)*, ∊\**(*t*), *represented in* (25), *which minimize the functional J over the control set U. Also, there exists an adjoint system of* 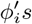 *such that the optimal medication,isolation and quarantine* (*susceptible*) *are characterized as* (24)*.Moreover, The adjoint system* (24) *satisfies the transversality conditions* {*ϕ_i_*(*T*) = 0, *i* = 1, 2,…, 10}.

### 8.2 Control Strategies and Comparison

We determine the optimal quarantine (*ϵ*(*t*) and isolation *τ*(*t*)) strategies when medication is available, we also consider the case when no medication is available. We would like to compare the quarantine and isolation strategies in both scenarios, this will help us understand the role medication may play in easing strict lock downs and isolation. We also determine the optimal medication strategy *α_I_* and the costs associated with implementing these strategies.

The Fig. 7(a) shows that there is a reduction in cost when medication is used in addition to quarantine and isolation, this may appear counter intuitive at first, but we note that the cost includes the cost of disease burden in addition to the cost associated with applying the control strategy itself.

**Figure 7:**
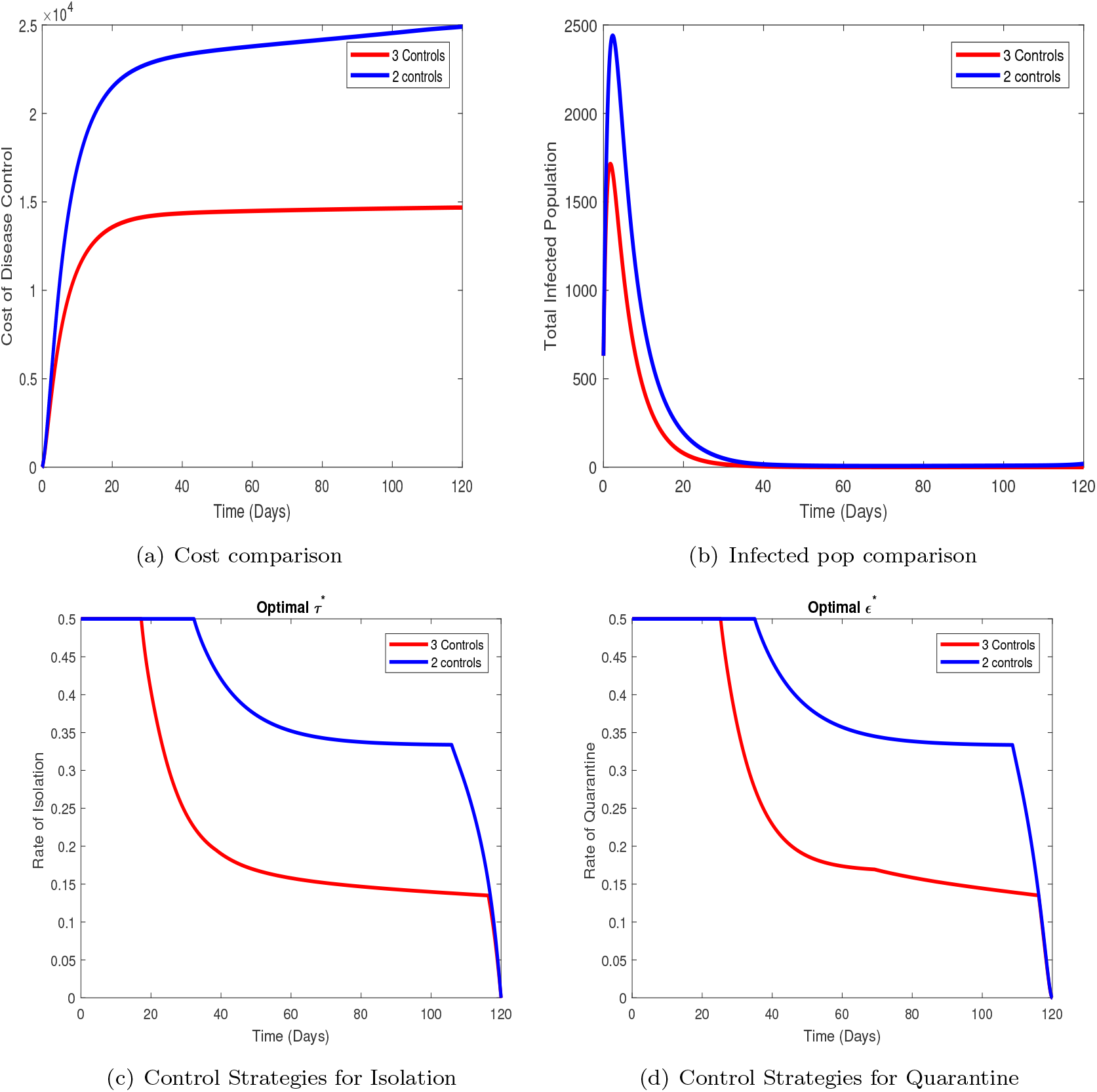
Optimal control comparison

This idea is made clearer by Fig7(b) in which the total optimal infected population over time is given under two and three controls. We see that the addition of medication as a control mechanism reduces the infected population, thereby reducing the cost which includes the disease burden. In Fig.7(c,d), isolation and quarantine strategies are shown in the presence and absence of medication. These graphs clearly indicate that using medication significantly reduces the quarantine and isolation rates needed to control the outbreak. This decrease is observed in both the time of maximum quarantine and isolation and the faster decrease in the rates over time.

We now consider the quarantine strategies for a variety of realistic contact rates *β* and for different medication efficacies *θ_M_*. The efficacy of medication is defined in terms of reducing the time it takes to recover from the disease once medication is started. As noted in the Introduction, several promising treatments are being evaluated, all of these in initial trials have reduced the symptoms and infectiousness. We note that for a higher contact rates and we require a more strict quarantine to be in place, this is reflected in the fact that the maximum rate of quarantine has to be maintained for a longer time and the drop in the quarantine rate is slower for higher contact rates. We also see that for lower medication efficacy, reflected in the average time for recovery after medication 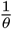, a stricter quarantine must be put in place and and eased at a a slower rate over a longer period of time. Our main finding is that the availability of medication will help in easing the strict quarantine in place, moreover the cost of implementing the control strategies (which includes the cost of disease burden) is also reduced with the availability and use of medication.

**Figure 8:**
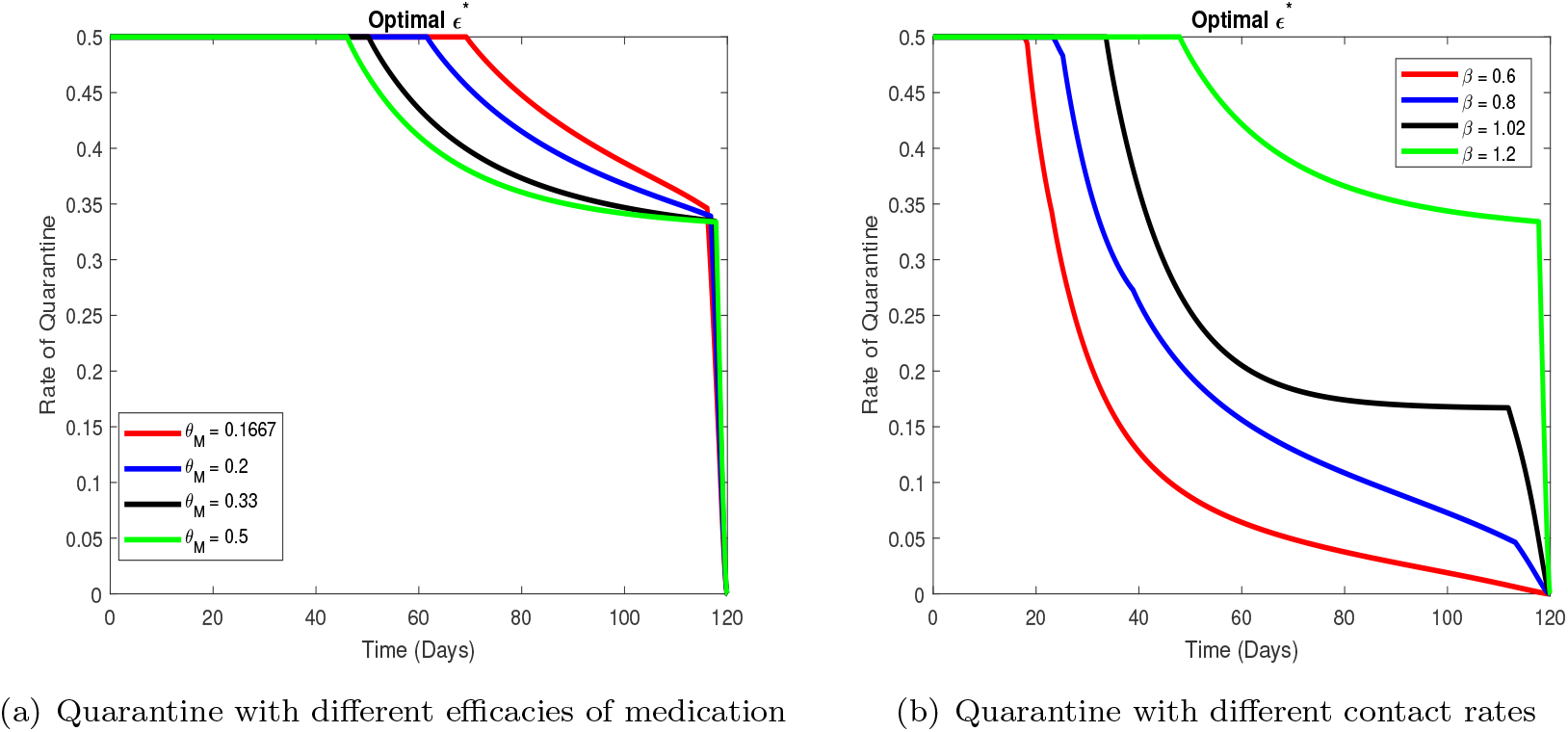
Optimal control comparison

## 9 Conclusion

The study considered a compartmental model for the transmission dynamics and control of COVID-19. We included quarantine, isolation as well as medication as possible control mechanisms for the disease. We look into how the availability of medication may affect the control strategies in place, specially the strict quarantine measures. We also studied how these measures depend on the rate and efficacy of the medication.

This model was an extension of an earlier model [24] incorporating a ‘treatment’ compartment. We established the existence of a threshold quantity *R*_0_ and showed that the disease dies out whenever *R*_0_ < 1 and is endemic in the population if *R*_0_ > 1, which basically means that in order to control the disease measures should be taken to reduce *R*_0_ to be less than 1. We also calculate the final size of the epidemic. We look at how *R*_0_ depends on the quarantine rate and the rate and efficacy of the medication. Our results show that using realistic values for the rate and efficacy of the treatment we still need to quarantine the susceptible population albeit at a smaller rate. We further study how the rate and efficacy affect the average time needed to stay in quarantine, our results show that with the availability of effective medication the average time in quarantine is significantly reduced. This leads us to conclude that once medication is available to a wider population the lock down measures can be eased. We estimated *R*_0_ for four different countries. India and Pakistan, where the curve is still rising, have shown very different COVID-19 transmission dynamics. The initial growth rates have been much slower with a much lower mortality rate as compared to China, Europe and North America. At this stage of the epidemic, *R*_0_ has been estimated to be 1.35 and 1.2 respectively. In Spain and Italy where the epidemic curve is now falling, after a strict lock down was imposed, *R*_0_ has been estimated to be 1.28 and 1,12 respectively. We further provided an estimate of the proportion of asymptomatic individuals in the population, this comes out to be around 25% which is in agreement with other studies.

We next looked at effective control measures. Using sensitivity analysis we concluded that *R*_0_ was most sensitive to the contact rate *β*, the rates of entering and leaving quarantine, *∊* and *ξ* respectively and the rate of medication of the infected population *α_i_*. Using this information we used optimal control theory to devise optimal quarantine, isolation and medication strategies. We also compared the strategies in the availability and non availability of medication. We observe that that quarantine is less severe when treatment is available. Moreover, we looked at the required quarantine rate for effectively controlling the disease, in the presence of medication for different realistic values of the contact rate and efficacy of the medication.

This study was motivated by the positive news regarding initial clinical trials of some plausible treatments for COVID-19. Thus far the only effective control measures were non pharmaceutical interventions including social distancing measures, quarantine and isolation, all of which have had a socio-economic cost. Many countries have started to relax these measures despite serious reservations expressed by the health authorities due to economic and political reasons. We have shown that with the availability of effective medication it may be possible to relax some of the measures while keeping the outbreak under control. However, our study also shows that along with treatment some level of social distancing will have to be maintained, at least until a vaccine is found. Quarantine and Isolation measures have resulted in bring down the value of *R*_0_ in Spain and Italy significantly, while in India and Pakistan where the outbreak is still in the growth phase, *R*_0_ values at the moment are low, however any premature relaxation in the lock down, specially till medication is widely available may result in much more severe outbreak.

## Data Availability

COVID-19 CURRENT REPORTING DATA BEING USED INTO THE MANUSCRIPT WHICH IS AVAILABLE HERE 
https://github.com/CSSEGISandData/COVID-19

## A Appendix

### A.1 Proof of Lemma 3.1

#### Proof

Adding the equations of model (2),this results in change in total population as

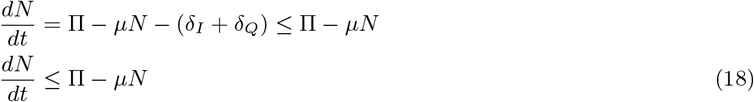

It follows that

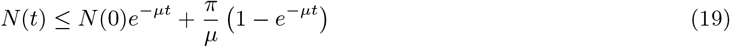

Thus model (2) solutions exists for given initial conditions, and are eventually bounded on every finite time interval. □

### A.2 Proof of Lemma 3.2

#### Proof

Using (18) and (19), it follows that as time approaches to infinity *t* → ∞, the population is bounded by the positive number so the set 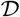

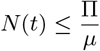

therefore, the set 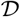 is positively invariant. □

### A.3 Proof of Lemma 4.2

#### Proof

Consider the Lyapunov function for model (2).

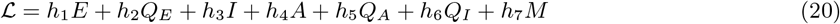

where

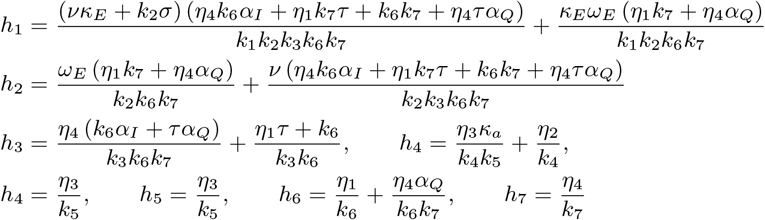

The Lyaponov derivative 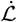 is given as

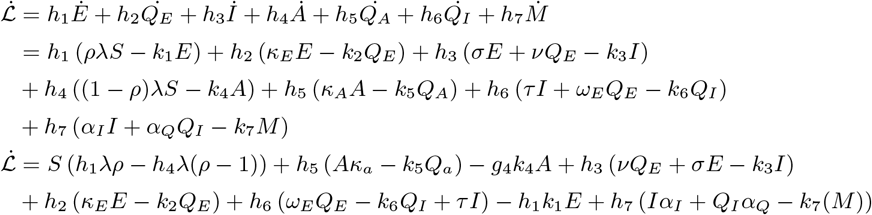

Since 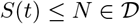, Thus

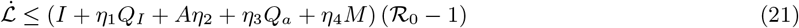

Thus, if 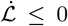 if 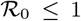 with 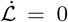 iff *E* = 0, *Q_E_* = 0, *I* = 0, *A* = 0, *Q_A_* = 0, *Q_I_* = 0 and *M* = 0. Additionally, the super compact set 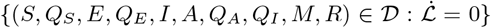 is the singleton 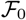. Thus, LaSalle Invariance Principle Theorem 6.4 [33] guaranties that every solution to the model (2) initial conditions from 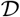 converge to DFE 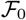 as *t* → ∞. Hence, (*E*, *Q_E_*, *I, A, Q_A_, Q_j_, M*) → (0, 0, 0, 0, 0, 0, 0) as *t* → ∞, It follows that 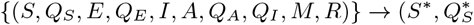 as *t* → ∞ for 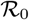. Hence, 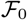 is 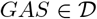 for 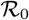.

### A.4 Proof of theorem 4.4

#### Proof

Since *m* = 1, *n* = 7 and *k* = 2, the above matrices can be written as

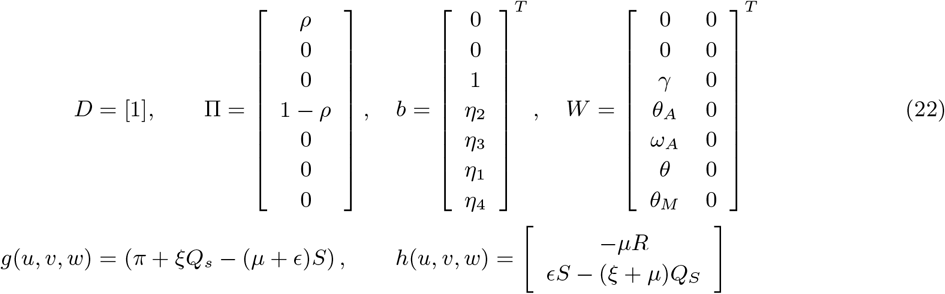

Consider the formulation of the m-dimentional row vector

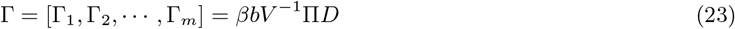

It follows from the theorem 2.1 from Arino et.al[34], 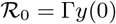. Also, the final size can be computed as

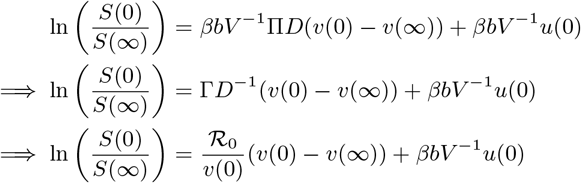

Thus, with the given initial conditions *u*(0), *v*(0), the final size of epidemic is represented by (8). □

### A.5 Proof of Theorem 4.5

#### Proof

Let 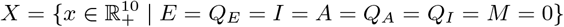 (that is, *X* is the disease-free subspace). Let *M* = *D* ⋂ *X*. Note that both *X*, as well as *M*, are positively invariant.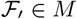 and it attracts all the trajectories in *X*. 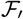 is asymptotically stable in *X*. Hence 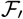 is isolated in *X*. Corollary 4.7 in [39] (where 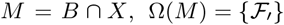, *T* = 1, *P*(1, *E*_0_) is 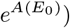, together with Proposition 4.1 and Lemma 3.1 in [39], imply that 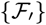 is uniformly weakly repelling. Then, from Theorem 8.17 in [40] we have that the semi-flow generated by (2.4) is uniformly weakly *ρ*-persistent. From the positive invariant of 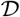, we have that (2.4) is point dissipative. Then, according to Theorem 2.28 in [40], there exists a compact attractor of points for the model (2). This, together with uniformly weakly *ρ*-persistent imply (11).

### A.6 Proof of Theorem 8.1

#### Proof

The Hamiltonian can be written as

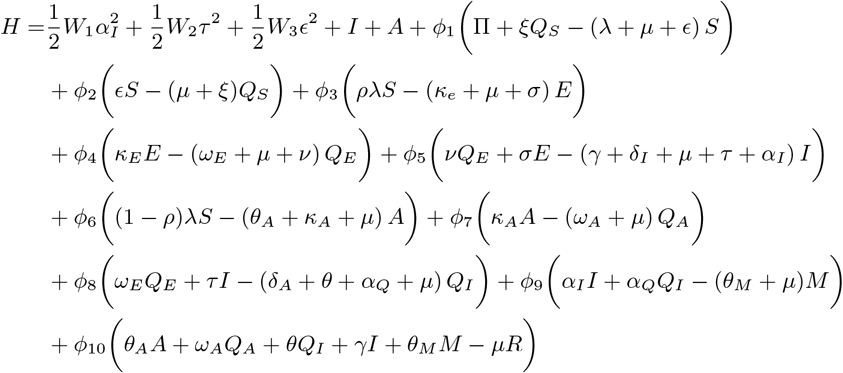

One can find that the Integrand *J*(·) is convex with respect to the control variables *α_I_*, *τ* and *ϵ*. By lemma (3.1), the state model (2) solutions are bound above 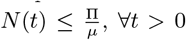.Moreover, The model follows Lipschitz property to the state variables. The convexity of the integrand *J*, boundedness of solutions of the state model and Lipschitz property guarantee us the optimal values of the control variables over the set U solutions [35]. Hence, establishing the existence of the control variables. The adjoint system is a acquired by means of Pontryain’s Maximum principle conditions

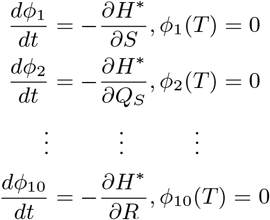

The adjoint system is given as

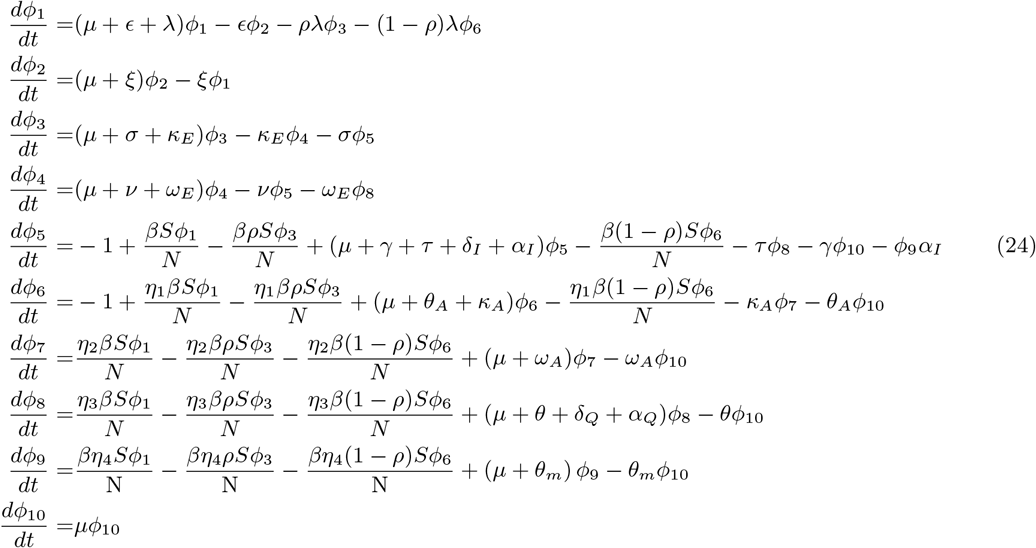

The optimality condition gives:

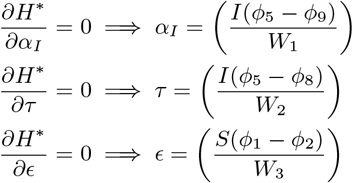

Since the control are bounded in set in *U*, setting the max limits to the controls *α_I_*, *τ* and *ϵ* by ζ_1_, ζ_2_ and ζ_3_ respectively. The optimal controls becomes:

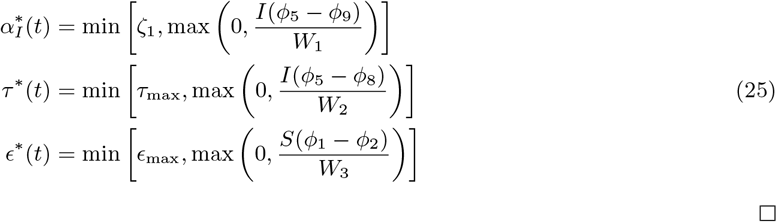

□

The uniqueness of optimal controls followed from the uniqueness of the optimal uniqueness of the optimality systems (state and adjoint).

## B Tables

**Table 3:**
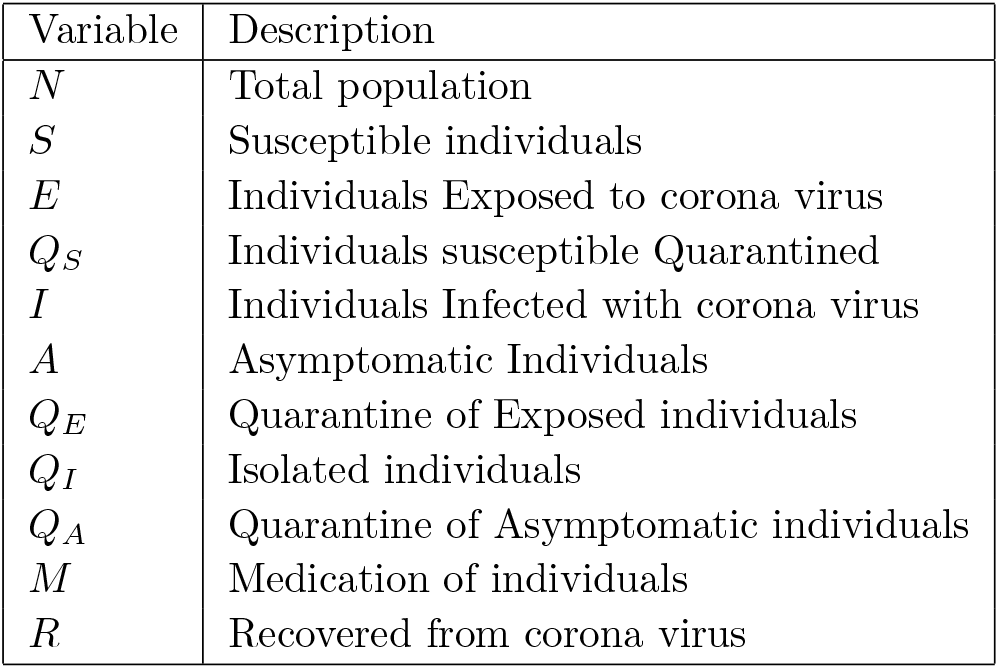
Description of the variables of the model

| Variable | Description |
| --- | --- |
| $N$ | Total population |
| $S$ | Susceptible individuals |
| $E$ | Individuals Exposed to corona virus |
| $Q_S$ | Individuals susceptible Quarantined |
| $I$ | Individuals Infected with corona virus |
| $A$ | Asymptomatic Individuals |
| $Q_E$ | Quarantine of Exposed individuals |
| $Q_I$ | Isolated individuals |
| $Q_A$ | Quarantine of Asymptomatic individuals |
| $M$ | Medication of individuals |
| $R$ | Recovered from corona virus |

**Table 4:** Description of the parameters of the model

| Parameters | Description | Values |  |
| --- | --- | --- | --- |
| $\Pi$ | Recruitment rate of humans | 100 | Assumed |
| $\mu$ | Natural death rate of humans | 60 years | Assumed |
| $\delta_I$ | Disease-induced death rate individuals | $0.065 \text{ day}^{-1}$ | Estimated |
| $\delta_Q$ | Disease-induced death rate of isolated individuals | $0.04 \text{ day}^{-1}$ | Assumed |
| $\nu$ | Rate of infectious of Exposed Quarantined individuals | 0.4 | Assumed |
| $\frac{1}{\sigma}$ | Incubation rate of susceptible individuals | 3 – 5 days | [23, 38] |
| $\frac{1}{\gamma}$ | Recovery rate of infected individuals | 10 days | [23, 38] |
| $\beta$ | Effective contact rate | 0.8 – 1.5 | [23] |
| $\rho$ | Probability of getting exposed to covid-19 | 40% = 0.4 | [23] |
| $\tau$ | Isolation rate of infected individuals | 0.2 | [23] |
| $\alpha_I$ | medication rate of infected individuals | 0.2 | Assumed |
| $\kappa_E$ | Quarantine rate of Exposed individuals | 0.4 | Assumed |
| $\kappa_A$ | Quarantine rate of Asymptomatic individuals | 0.4 | Assumed |
| $\omega_E$ | Isolation rate of Exposed Quarantined individuals | 0.1 | Assumed |
| $\omega_A$ | Recovery rate of Exposed Asymptomatic individuals | 0.1 | [23, 38] |
| $\theta_A$ | Recovery rate of Asymptomatic individuals | 0.3 | [23, 38] |
| $\theta$ | Recovery rate of Isolated individuals | 0.3 | [23, 38] |
| $\theta_M$ | Recovery rate of medicated individuals | 0.4 | Assumed |
| $\epsilon$ | susceptible Quarantine rate of Susceptible individuals | 0.5 | Assumed |
| $\xi$ | Waning rate of susceptible quarantined individuals to susceptible class | 0.4 | Assumed |
| $\eta_1$ | Modification parameter for relative infectiousness of Isolated individuals | 0.2 | Assumed |
| $\eta_2$ | Modification parameter for relative infectiousness of asymptomatic individuals | 0.5 | [23] |
| $\eta_3$ | Modification parameter for relative infectiousness of asymptomatic quarantined | 0.2 | Assumed |
| $\eta_4$ | Modification parameter for relative infectiousness of medication | 0.2 | Assumed |

## References

[1] Centers for Disease Control and Prevention. 2020. Coronavirus Disease 2019 (COVID-19). [online] Available at: https://www.cdc.gov/coronavirus/2019-ncov/ [Accessed 25 April 2020].

[2] Who. int. 2020. Coronavirus. [online] Available at: https://www.who.int/health-topics/coronavirus [Accessed 25 April 2020].

[3] Worldometers. info. 2020. Coronavirus Update (Live): 2,874,022 Cases And 200,790 Deaths From COVID-19 Virus Pandemic - Worldometer. [online] Available at: https://www.worldometers.info/coronavirus/ [Accessed 25 April 2020].

[4] Oie. int. 2020. Questions And Answers On The COVID-19: OIE - World Organisation For Animal Health. [online] Available at: https://www.oie.int/scientific-expertise/specific-information-and-recommendations/questions-and-answers-on-2019novel-coronavirus [Accessed 25 April 2020].

[5] Hu B, Ge X, Wang L, Shi Z. Bat origin of human coronaviruses. Virol J. 2015;12(1). doi:10.1186/s12985-015-0422-1

[6] Huang. C et al. Clinical features of patients infected with 2019 novel coronavirus in Wuhan, China. Lancet 2020; 395: 497–506

[7] Nicola. M, Alsafi. Z, Sohrabi. C, Kerwan. A, Al-Jabir. A, Iosifidis. C, Agha. M, Agha. R. The Socio-Economic Implications of the Coronavirus and COVID-19 Pandemic: A Review. International Journal of Surgery. https://doi.org/10.1016/j.ijsu.2020.04.018.

[8] Sallard, E., Lescure, F., Yazdanpanah, Y., Mentre, F. and Peiffer-Smadja, N., 2020. Type 1 interferons as a potential treatment against COVID-19. Antiviral Research, 178, p.104791.

[9] Beigel, J., Tomashek, K., Dodd, L., Mehta, A., Zingman, B., Kalil, A., Hohmann, E., Chu, H., Luetkemeyer, A., Kline, S., Lopez de Castilla, D., Finberg, R., Dierberg, K., Tapson, V., Hsieh, L., Patterson, T., Paredes, R., Sweeney, D., Short, W., Touloumi, G., Lye, D., Ohmagari, N., Oh, M., Ruiz-Palacios, G., Benfield, T., Fatkenheuer, G., Kortepeter, M., Atmar, R., Creech, C., Lundgren, J., Babiker, A., Pett, S., Neaton, J., Burgess, T., Bonnett, T., Green, M., Makowski, M., Osinusi, A., Nayak, S. and Lane, H., 2020. Remdesivir for the Treatment of Covid-19 — Preliminary Report. New England Journal of Medicine. DOI: 10.1056/NEJMoa2007764

[10] Mehra, M., Desai, S., Ruschitzka, F. and Patel, A. Hydroxychloroquine or chloroquine with or without a macrolide for treatment of COVID-19: a multinational registry analysis. The Lancet 2020. https://doi.org/10.1016/S0140-6736(20)31180-6

[11] 2. Wang. Y, Zhang. D, Du. G et al. Remdesivir in adults with severe COVID-19: a randomised, doubleblind, placebo-controlled, multicentre trial. The Lancet. 2020;395(10236):1569–1578. doi:10.1016/s0140-6736(20)31022-9

[12] Hung, I., Lung, K., Tso, E., Liu, R., Chung, T., Chu, M., Ng, Y., Lo, J., Chan, J., Tam, A., Shum, H., Chan, V., Wu, A., Sin, K., Leung, W., Law, W., Lung, D., Sin, S., Yeung, P., Yip, C., Zhang, R., Fung, A., Yan, E., Leung, K., Ip, J., Chu, A., Chan, W., Ng, A., Lee, R., Fung, K., Yeung, A., Wu, T., Chan, J., Yan, W., Chan, W., Chan, J., Lie, A., Tsang, O., Cheng, V., Que, T., Lau, C., Chan, K., To, K. and Yuen, K., 2020. Triple combination of interferon beta-1b, lopinavir-ritonavir, and ribavirin in the treatment of patients admitted to hospital with COVID-19: an open-label, randomised, phase 2 trial. The Lancet,.

[13] Grein, J., Ohmagari, N., Shin, D., Diaz, G., Asperges, E., Castagna, A., Feldt, T., Green, G., Green, M., Lescure, F., Nicastri, E., Oda, R., Yo, K., Quiros-Roldan, E., Studemeister, A., Redinski, J., Ahmed, S., Bernett, J., Chelliah, D., Chen, D., Chihara, S., Cohen, S., Cunningham, J., D’Arminio Monforte, A., Ismail, S., Kato, H., Lapadula, G., L’Her, E., Maeno, T., Majumder, S., Massari, M., Mora-Rillo, M., Mutoh, Y., Nguyen, D., Verweij, E., Zoufaly, A., Osinusi, A., DeZure, A., Zhao, Y., Zhong, L., Chokkalingam, A., Elboudwarej, E., Telep, L., Timbs, L., Henne, I., Sellers, S., Cao, H., Tan, S., Winterbourne, L., Desai, P., Mera, R., Gaggar, A., Myers, R., Brainard, D., Childs, R. and Flanigan, T., 2020. Compassionate Use of Remdesivir for Patients with Severe Covid-19. New England Journal of Medicine,.

[14] Yang, J., Zheng, Y., Gou, X., Pu, K., Chen, Z., Guo, Q., Ji, R., Wang, H., Wang, Y. and Zhou, Y. Prevalence of comorbidities and its effects in coronavirus disease 2019 patient: A systematic review and meta-analysis. International Journal of Infectious Diseases, 94, pp.91 — 95 2020

[15] https://www.nih.gov/news-events/news-releases/nih-begins-clinical-trial-hydroxychloroquine-azithromycin-treat-covid-19

[16] Zhao S, Lin Q, Ran J et al. Preliminary estimation of the basic reproduction number of novel coronavirus (2019-nCoV) in China, from 2019 to 2020: A data-driven analysis in the early phase of the outbreak. International Journal of Infectious Diseases. 2020;92:214–217. doi:10.1016/j.ijid.2020.01.050

[17] 2. Zhao S, Lin Q, Ran J et al. Preliminary estimation of the basic reproduction number of novel coronavirus (2019-nCoV) in China, from 2019 to 2020: A data-driven analysis in the early phase of the outbreak. International Journal of Infectious Diseases. 2020;92:214–217. doi:10.1016/j.ijid.2020.01.050

[18] Kucharski A, Russell T, Diamond C et al. Early dynamics of transmission and control of COVID-19: a mathematical modelling study. The Lancet Infectious Diseases. 2020. doi:10.1016/s1473-3099(20)30144-4

[19] Chen T, Rui J, Wang Q, Zhao Z, Cui J, Yin L. A mathematical model for simulating the phase-based transmissibility of a novel coronavirus. Infect Dis Poverty. 2020;9(1). doi:10.1186/s40249-020-00640-3

[20] Lin Q, Zhao S, Gao D et al. A conceptual model for the coronavirus disease 2019 (COVID-19) outbreak in Wuhan, China with individual reaction and governmental action. International Journal of Infectious Diseases. 2020;93:211–216. doi:10.1016/j.ijid.2020.02.058

[21] 8. Bhatnagar T, Mandal S, Arinaminpathy N et al. Prudent public health intervention strategies to control the coronavirus disease 2019 transmission in India: A mathematical model-based approach. Indian Journal of Medical Research. 2020;0(0):0. doi:10.4103/ijmr.ijmr50420

[22] Aslan. H.I et al. Modeling COVID-19: Forecasting and analyzing the dynamics of the outbreak in Hubei and Turkey. Preprint doi: https://doi.org/10.1101/2020.04.11.20061952

[23] Eikenberry S, Mancuso M, Iboi E, Phan T, Eikenberry K, Kuang Y, Kostelich E. and Gumel A. To mask or not to mask: Modeling the potential for face mask use by the general public to curtail the COVID-19 pandemic. Infectious Disease Modelling, 5, pp.293–308 2020.

[24] Ali, M., Shah, S., Imran, M. and Khan, A., 2020. The Role of Asymptomatic Class, Quarantine and Isolation in the transmission of COVID-19. Journal of Biological Dynamics,.(Accepted)

[25] Lee, S., Chowell, G. and Castillo-Chavez, C., 2010. Optimal control for pandemic influenza: The role of limited antiviral treatment and isolation. Journal of Theoretical Biology, 265(2), pp.136–150.

[26] Jia, W., Weng, J., Fang, C. and Li, Y., 2019. A dynamic model and some strategies on how to prevent and control hepatitis c in mainland China. BMC Infectious Diseases, 19(1).

[27] Granich, R., Gilks, C., Dye, C., De Cock, K. and Williams, B., 2009. Universal voluntary HIV testing with immediate antiretroviral therapy as a strategy for elimination of HIV transmission: a mathematical model. The Lancet, 373(9657), pp.48–57.

[28] Sharomi, O., Podder, C. and Gumel, A., 2008. Mathematical analysis of the transmission dynamics of HIV/TB coinfection in the presence of treatment. Mathematical Biosciences and Engineering, 5(1), pp.145–174.

[29] Imran, M., Malik, T., Ansari, A. and Khan, A., 2016. Mathematical analysis of swine influenza epidemic model with optimal control. Japan Journal of Industrial and Applied Mathematics, 33(1), pp.269–296.

[30] Pang L, Liu S, Zhang X, Tian T. and Zhao Z. Transission Dynamics and Control Strategies of COVID-19 in Wihan China. Journal of Biological Systems, pp.1–18 2020.

[31] Perkins A. and Espana, G. Optimal control of the COVID-19 pandemic with non-pharmaceutical interventions. 10.1101/2020.04.22.20076018 (Preprint)

[32] P. van den Driessche, and J. Watmough. “Reproduction Numbers and Sub-Threshold Endemic Equilibria for Compartmental Models of Disease Transmission,” Mathematical Biosciences, Vol. 180, pp. 29 — 48, 2002.

[33] LaSalle, J.P. (1976). The Stability of Dynamical Systems. Regional Conference Series in Applied Mathematics, SIAM, Philadelphia.

[34] Arino J., Brauer F., Van Den Driessche P., Watmough J., Wu J. “A final size relation for epidemic models” Math. Biosci. Eng., 4 (2) (2007), p. 159

[35] W. Fleming and R. Rishel, Deterministic and Stochastic Optimal Control (Springer-Verlag, Berlin, 1975).

[36] D. Kirschner, S. Lenhart and S. Serbin, Optimal control of the chemotherapy of HIV, J. Math. Biol. 35(7) (1997) 775–792.

[37] L. S. Pontryagin and V. G. Boltyanskii, The Mathematical Theory of Optimal Processes (Golden and Breach Science Publishers, 1980).

[38] Ferguson, N., Laydon, D., Nedjati Gilani, G., Imai, N., Ainslie, K., Baguelin, M., Bhatia, S., Boonyasiri, A., Cucunuba Perez, Z., Cuomo-Dannenburg, G., Dighe, A., Dorigatti, I., Fu, H., Gaythorpe, K., Green, W., Hamlet, A., Hinsley, W., Okell, L., Van Elsland, S., Thompson, H., Verity, R., Volz, E., Wang, H., Wang, Y., Walker, P., Walters, C., Winskill, P., Whittaker, C., Donnelly, C., Riley, S. and Ghani, A., 2020. Report 9: Impact Of Non-Pharmaceutical Interventions (Npis) To Reduce COVID19 Mortality And Healthcare Demand. [online] Hdl.handle.net. Available at: http://hdl.handle.net/10044/1/77482 [Accessed 2 May 2020].

[39] P. L. Salceanu, Robust uniform persistence in discrete and continuous dynamical systems using Lyapunov Exponents, Math. Biosci. Eng, 8(3), 2011, pp 807–825.

[40] H. L. Smith, H. Thieme, Dynamical Systems and Population Persistence, Graduate Studies in Mathematics, Amer. Math. Soc, 118, 2011.

[41] Data.humdata.org. 2020. Novel Coronavirus (COVID-19) Cases Data - Humanitarian Data Exchange. [online] Available at: https://data.humdata.org/dataset/novel-coronavirus-2019-ncov-cases [Accessed 21 May 2020].

[42] A. Cintron-Arias, C. Castillo-Chavez, L.M.A. Bettencourt, A.L. Lloyd, and H.T. Banks, The estimation of the effective reproductive number from disease outbreak data, Math. Biosciences Eng. 6 (2009), pp. 261–282. doi:10.3934/mbe.2009.6.261

[43] B. Efron and R.J. Tibshirani, An Introduction to the Bootstrap, Chapman & Hall/CRC, Boca Raton, FL, 1993.

